# Dual-Energy Pulsed Field and Radiofrequency Ablation With a Contact Force–Sensing Focal Catheter for Ventricular Arrhythmias: Initial Clinical Experience

**DOI:** 10.64898/2026.10.19.26365958

**Authors:** Angeliki Darma, Said-Elias Waezsada, Nadica Trajkovska, Moneeb Khalaph, Hassan El-shirbiny, Stjepan Jurisic, Philipp Lucas, Ersan Akkaya, Max Moersdorf, Thomas Fink, Vanessa Sciacca, Philipp Sommer, Christian Sohns

**Affiliations:** Department of Electrophysiology, Heart and Diabetes Center NRW, Ruhr University Bochum, Bad Oeynhausen, Germany; Cardiology Department, Faculty of Medicine, Kafr Elsheikh University, Kafr El Sheikh, Egypt; Department of Cardiology, University Hospital Zurich, Zurich, Switzerland

**Keywords:** pulsed field ablation, radiofrequency ablation, dual energy, ventricular tachycardia, premature ventricular contractions, structural heart disease

## Abstract

**Background:** Clinical data on ventricular ablation with a contact force–sensing focal catheter capable of delivering both radiofrequency (RF) and pulsed field (PF) energy are limited.

**Objective:** To describe the procedural characteristics, acute efficacy, and safety of dual-energy ablation with the Dual Energy THERMOCOOL SMARTTOUCH SF (DE-STSF) catheter in consecutive patients with ventricular arrhythmias.

**Methods:** This observational single-center study included all consecutive patients who underwent catheter ablation of premature ventricular contractions (PVCs) or ventricular tachycardia (VT) with the DE-STSF catheter between June and October 2026.

**Results:** Twenty-six patients were included (median age 65.5 [55.8–71.0] years; 77% male; median left ventricular ejection fraction 44.0% [30.0–53.8]). Eighteen underwent VT ablation, including 10 with electrical storm or incessant VT and 6 with prior VT ablation; 8 underwent PVC ablation. All patients received RF followed by PF consolidation. Noninducibility of any sustained VT or ventricular fibrillation was achieved in 16/18 VT patients (89%), and acute PVC elimination in all 8 PVC patients. No major procedure-related complications were documented during hospitalization. During a median follow-up of 68 [47–82] days, VT recurred in 2 patients; of whom one patient underwent repeat endocardial and epicardial ablation. No PVC recurrences were documented.

**Conclusion:** In this initial clinical experience, sequential RF and PF ablation using a single contact force–sensing catheter proved feasible for the treatment of ventricular arrhythmias, achieving high acute success with a favorable early safety profile. Whether PF consolidation confers incremental benefit beyond RF ablation alone warrants prospective comparative evaluation.

**Clinical Perspective:** *What is Known:*

- Radiofrequency ablation may be limited by ventricular wall thickness and complex scar, and focal pulsed field and dual-energy approaches have been explored in early clinical studies.
- Clinical evidence is specific to the catheter platform and energy-delivery protocol, and the incremental benefit of sequential RF and PF delivery remains uncertain.

*What the Study Adds:*

- A sequential RF-first and PF-consolidation workflow with the contact force–sensing DE-STSF catheter was feasible in 26 consecutive patients undergoing VT or PVC ablation, including selected epicardial applications.
- Acute noninducibility was achieved in 16 of 18 VT patients and acute PVC elimination in all 8 PVC patients; no major in-hospital procedure-related complications were documented, although one transient LV lead parameter change occurred.
- These encouraging initial results support further prospective evaluation of the DE-STSF catheter for ventricular arrhythmia ablation, with a focus on optimizing energy delivery and assessing long-term efficacy and safety.

## Introduction

Catheter ablation is an established treatment for recurrent ventricular tachycardia (VT) in patients with structural heart disease and for symptomatic or cardiomyopathy-inducing premature ventricular contractions (PVCs). (1,2) Freedom from VT recurrence after ablation is associated with improved survival, (3) yet long-term outcomes remain unsatisfactory, particularly in patients with nonischemic cardiomyopathy. A central limitation of radiofrequency (RF) energy is its restricted and variable lesion depth in thick, fibrotic, or fat-insulated myocardium; in addition, RF delivery near the conduction system, coronary arteries, or the phrenic nerve is constrained by the risk of collateral thermal injury.

Pulsed field ablation (PFA) creates lesions by irreversible electroporation and has rapidly transformed atrial fibrillation ablation, with randomized and large-scale registry data supporting its efficacy and safety profile. (4,5,6) Its relative myocardial selectivity and the potential to create lesions less dependent on tissue heating make PFA an attractive option for ventricular substrate. Preclinical work has shown that focal PF lesions in ventricular myocardium can reach clinically relevant depths, which increase with repeated applications, (7) while dual-energy focal catheters allow sequential delivery of RF and PF energy within a single procedural workflow. (8) In a preclinical DE-STSF model, sequential, colocalized RF and PF delivery increased lesion dimensions compared with either energy alone; whether this translates into clinical benefit in ventricular scar is unknown. (9) At the same time, ventricular PFA raises specific concerns, including coronary spasm, hemolysis, and device damage during energy delivery.

Clinical experience with ventricular PFA includes early clinical series and a multicenter registry using different focal and dual-energy platforms. (10,11,12,13) Evidence specific to individual catheters and ventricular energy-delivery protocols remains limited. The Dual Energy THERMOCOOL SMARTTOUCH SF (DE-STSF) catheter combines contact force sensing, irrigation, and full integration into a widely used three-dimensional mapping system with the ability to deliver RF and PF energy from the same tip. The platform received CE mark approval based on data in paroxysmal atrial fibrillation, (14,15) and its first use for a ventricular arrhythmia was reported only recently. (16,17,18) Prospective studies in PF ablation for ventricular arrhythmias are ongoing. (19,20)

We therefore report our initial consecutive clinical experience with dual-energy ablation using the DE-STSF catheter in patients undergoing ablation of VT and PVCs, focusing on procedural characteristics, acute efficacy, and safety.

## Methods

### Study design and population

This prospective, observational, single-center study included all consecutive patients who underwent catheter ablation of VT or PVCs with the DE-STSF catheter (Johnson & Johnson MedTech, Irvine, CA, USA) at the Heart and Diabetes Center North Rhine-Westphalia between June and October 2026. During this period, 27 ventricular procedures were performed with this system at our institution, comprising the index procedures of 26 patients and one repeat procedure in a patient with VT recurrence; only index procedures were included in the analysis, and the repeat procedure was not counted as a separate patient. Indications for ablation followed current guidelines. (1,2) The study complied with the Declaration of Helsinki and was approved by the local ethics committee.

Baseline data included demographics, underlying substrate (ischemic cardiomyopathy [ICM], nonischemic cardiomyopathy [NICM], arrhythmogenic right ventricular cardiomyopathy [ARVC], or idiopathic PVC without known cardiomyopathy), NYHA class, comorbidities, antiarrhythmic drug therapy, number of prior ablation procedures, left ventricular ejection fraction (LVEF), and the presence of electrical storm or incessant VT.

### Electrophysiological study and ablation strategy

All patients provided written informed consent prior to the procedure. Procedures were performed under general anesthesia for VT ablation and conscious sedation for PVC ablation. Intravenous heparin was administered to maintain an activated clotting time of at least 300 seconds during left ventricular mapping; otherwise, a prophylactic heparin bolus was administered at the beginning of the procedure.

Vascular access was obtained via the femoral veins: two 7F sheaths in the left femoral vein and an 8.5F steerable sheath (Carto Vizigo, Johnson & Johnson MedTech, Irvine, CA, USA, or Agilis, Abbott, Chicago, IL, USA) in the right femoral vein. A decapolar catheter (Inquiry, Abbott) was positioned in the distal coronary sinus, and a quadripolar catheter (Supreme, Abbott) was placed at the right ventricular apex. Right ventricular mapping was performed via the right femoral vein. Left ventricular mapping was carried out using a retrograde aortic and/or transseptal approach, with transseptal puncture performed in the anterior-inferior fossa ovalis under fluoroscopic guidance. In patients undergoing VT ablation, the standardized institutional workflow included right and left ventricular voltage mapping, programmed ventricular stimulation for VT induction, activation mapping of hemodynamically tolerated VT when feasible, ablation during tachycardia to assess termination, and subsequent substrate modification. PVCs were targeted using activation and pace mapping.

Electroanatomical mapping was performed with the CARTO 3 system (Johnson & Johnson MedTech). In VT procedures, high-density mapping was performed with the OPTRELL mapping catheter (Johnson & Johnson MedTech); in PVC procedures, mapping was performed with the DE-STSF ablation catheter itself. Low-voltage areas were defined as bipolar voltages between 0.1 and 1.5 mV. Epicardial access was attempted in 4 patients using a subxiphoid puncture according to the technique described by Sosa et al. (21). After successful access, a long steerable epicardial sheath (Agilis Epi, Abbott) was placed; at the end of the procedure, the sheath was exchanged for a pigtail catheter that remained in situ until the following day.

Programmed ventricular stimulation was performed from the right ventricular apex using a stepwise protocol with drive-train cycle lengths from 500 to 330 ms and up to four extrastimuli, both for VT induction and for repeat testing at the end of the procedure. The primary procedural endpoint for VT ablation was elimination of the clinical VT and noninducibility of any sustained VT or ventricular fibrillation; for PVC procedures, acute success was defined as the absence of the clinical PVC during a waiting period of 30 minutes after the last ablation lesion.

### Dual-energy ablation system and energy delivery

All ablations were performed with the Dual Energy THERMOCOOL SMARTTOUCH SF (DE-STSF) catheter (Johnson & Johnson MedTech), a 7.5F open-irrigated, contact force–sensing focal catheter with a 3.5-mm tip electrode connected to the TRUPULSE generator, which allows the operator to toggle between irrigated RF energy and biphasic, monopolar, microsecond PF energy. The system is fully integrated into CARTO 3, permitting automated lesion tagging for both energy modalities.

RF energy was delivered in power-controlled mode at 40 to 50 W, depending on the individual case and the ablation site, with irrigation settings according to the manufacturer’s recommendations. PF energy was delivered with the waveform parameters predefined by the TRUPULSE generator, which are not adjustable by the operator, with an irrigation flow of 4 ml/min, a contact force working range of 5 to 40 g, and applications of up to 28 seconds separated by 1-second intervals, as specified in the instructions for use. The routine dual-energy workflow consisted of RF delivery first, with PVC suppression or VT termination occurring during RF delivery, followed by PF applications for lesion consolidation. This sequence was chosen to avoid relying on acute arrhythmia suppression during PF delivery alone, which could reflect transient myocardial stunning rather than durable ablation. PF was used routinely for consolidation rather than as a bailout after unsuccessful RF delivery. One PF application was defined as one complete delivery cycle, represented by one PF lesion tag in CARTO 3. Repeated PF applications at the same site were delivered to achieve a target PF index of 600–700. A preclinical study using DE-STSF and TRUPULSE 2 evaluated index-guided ventricular lesion formation, including a target value of 650. (22) This was an operator-selected target rather than a clinically validated ventricular threshold for DE-STSF. All patients received both RF and PF lesions; at sites in the left ventricular outflow tract or where close proximity to a coronary artery was anticipated, only RF energy was delivered. Coronary angiography was not routinely performed, and prophylactic nitroglycerin was not routinely administered; in one patient with an LV summit PVC, RF and PF energy were delivered within the distal coronary sinus after administration of nitroglycerin. Procedure duration was defined as the skin-to-skin time, the ablation phase duration as the interval from the first to the last ablation application, and the total energy delivery time as the cumulative duration of all RF and PF applications.

### Safety endpoints and follow-up

The safety endpoint comprised major procedure-related complications during the index hospitalization, including death, stroke or transient ischemic attack, pericardial tamponade, vascular complications requiring intervention, atrioventricular block requiring pacing, coronary injury or spasm, acute kidney injury or clinically relevant hemolysis, and device malfunction.

Follow-up was defined as the period from the index ablation procedure to the most recent documented clinical evaluation. It included review of inpatient and outpatient medical records, ICD interrogations, remote monitoring, and communication with referring cardiologists and primary care physicians. VT recurrence was defined as any documented sustained ventricular arrhythmia, including VT or ventricular fibrillation, regardless of termination by antitachycardia pacing or appropriate ICD shock. Early recurrence was defined as VT recurrence during the index hospitalization. Electrical storm was defined as three or more episodes of sustained ventricular arrhythmia within 24 hours. In patients with a cardiac implantable electronic device (CIED), a complete device interrogation, including sensing, pacing thresholds, and lead impedances, was performed before and after the procedure and repeated on the following day if abnormalities were detected; tachyarrhythmia therapies were deactivated during the procedure. For PVC procedures, recurrence of the clinical PVC was assessed by continuous telemetry during the index hospitalization and by ambulatory Holter monitoring thereafter.

### Statistical analysis

Given the sample size, analyses were descriptive. Continuous variables are reported as median [interquartile range] and categorical variables as n (%). Results are presented for the overall cohort and separately for VT and PVC procedures; no formal between-group comparisons were performed because the groups differ fundamentally in substrate and procedural goals. Fluoroscopy time was recorded in minutes:seconds and converted to decimal minutes. Follow-up duration was calculated from the procedure date to the most recent documented clinical evaluation. Analyses were performed with SPSS version 24.0 (IBM Corp., Chicago, Illinois, USA) and Python (version 3.12.3).

## Results

### Patient characteristics

Twenty-six patients were included (median age 65.5 [55.8–71.0] years; 20 [77%] male; median LVEF 44.0% [30.0–53.8]). Eighteen patients (69%) underwent VT ablation and 8 (31%) PVC ablation (Table 1).

**Table 1.** Baseline characteristics.

| Characteristic | All (n = 26) | VT (n = 18) | PVC (n = 8) |
| --- | --- | --- | --- |
| Age, years | 65.5 [55.8–71.0] | 67.0 [60.2–72.0] | 55.0 [44.8–67.2] |
| Male sex | 20 (76.9) | 18 (100) | 2 (25.0) |
| Body mass index, kg/m<sup>2</sup> | 29.0 [27.4–32.3] | 28.3 [27.4–31.3] | 33.6 [27.8–39.9] |
| **Underlying substrate** |  |  |  |
| Ischemic cardiomyopathy | 9 (34.6) | 8 (44.4) | 1 (12.5) |
| Nonischemic cardiomyopathy | 7 (26.9) | 6 (33.3) | 1 (12.5) |
| ARVC | 4 (15.4) | 4 (22.2) | 0 |
| Idiopathic | 6 (23.1) | 0 | 6 (75.0) |
| LVEF, % | 44.0 [30.0–53.8] | 38.0 [30.0–50.0] | 54.0 [43.8–60.0] |
| LVEF ≤35% | 9 (34.6) | 9 (50.0) | 0 |
| NYHA class II / III | 25 / 1 | 17 / 1 | 8 / 0 |
| Arterial hypertension | 14 (53.8) | 11 (61.1) | 3 (37.5) |
| Diabetes mellitus type 2 | 2 (7.7) | 2 (11.1) | 0 |
| Chronic kidney disease | 6 (23.1) | 5 (27.8) | 1 (12.5) |
| Prior stroke/TIA | 1 (3.8) | 1 (5.6) | 0 |
| Atrial fibrillation | 8 (30.8) | 7 (38.9) | 1 (12.5) |
| **Antiarrhythmic therapy** |  |  |  |
| β-blocker only | 13 (50.0) | 6 (33.3) | 7 (87.5) |
| Amiodarone | 10 (38.5) | 10 (55.6) | 0 |
| Other | 2 (7.7) | 2 (11.1) | 0 |
| None | 1 (3.8) | 0 | 1 (12.5) |
| Prior ablation | 7 (26.9) | 6 (33.3) | 1 (12.5) |
| Electrical storm / incessant VT | 10/18 (55.6) | 10 (55.6) | n.a. |
| Implanted ICD / CRT-D / S-ICD | 17 (65.4) | 17 (94.4) | 0 |
| Prior ATP | 16/18 (88.9) | 16 (88.9) | n.a. |
| Prior ICD shock | 15/18 (83.3) | 15 (83.3) | n.a. |

Values are median [interquartile range] or n (%). ARVC = arrhythmogenic right ventricular cardiomyopathy; ATP = antitachycardia pacing; CRT-D = cardiac resynchronization therapy defibrillator; ICD = implantable cardioverter-defibrillator; S-ICD = subcutaneous ICD; LVEF = left ventricular ejection fraction; NYHA = New York Heart Association; PVC = premature ventricular contraction; TIA = transient ischemic attack; VT = ventricular tachycardia. n.a. = not applicable.

Patients undergoing VT ablation were all male, with a median age of 67.0 [60.2–72.0] years and a median LVEF of 38.0% [30.0–50.0]; 9 (50%) had an LVEF ≤35%. The underlying substrate was ICM in 8 (44%), NICM in 6 (33%), and ARVC in 4 (22%). The population was characterized by a high arrhythmic burden: 10 patients (56%) presented with electrical storm or incessant VT, 16 (89%) had received antitachycardia pacing, and 15 (83%) ICD shocks. Six patients (33%) had undergone at least one prior VT ablation (1 to 3 prior procedures), and 10 (56%) were treated with amiodarone.

Patients undergoing PVC ablation were younger (median 55.0 [44.8–67.2] years), predominantly female (6/8), and had a median LVEF of 54.0% [43.8–60.0]. Six patients had idiopathic PVCs without known cardiomyopathy, 1 had NICM, and 1 had ICM. PVC origins were the septal, posterior, and anterior right ventricular outflow tract (RVOT), the aortomitral continuity, the anterolateral papillary muscle, the basal anterolateral left ventricle (LV), and the LV summit (2 patients) (Table S2).

### Procedural characteristics

Combined PF and RF energy was delivered in all patients. Epicardial access was attempted in 4 patients and was successful in 3 (1 with ARVC and 2 with NICM), who underwent epicardial mapping and ablation; in 1 NICM patient, ablation was performed exclusively from the epicardium, as no endocardial scar was detected. One patient with ICM, an LVEF of 26%, and electrical storm underwent Impella CP-protected ablation. Ablation targets in VT patients involved the RV free wall and/or RVOT in all ARVC patients and the basal to apical LV in patients with ICM and NICM, including septal substrate in 11 patients (Table S1). Representative electroanatomical maps are shown in Figure 1.

**Figure 1.**
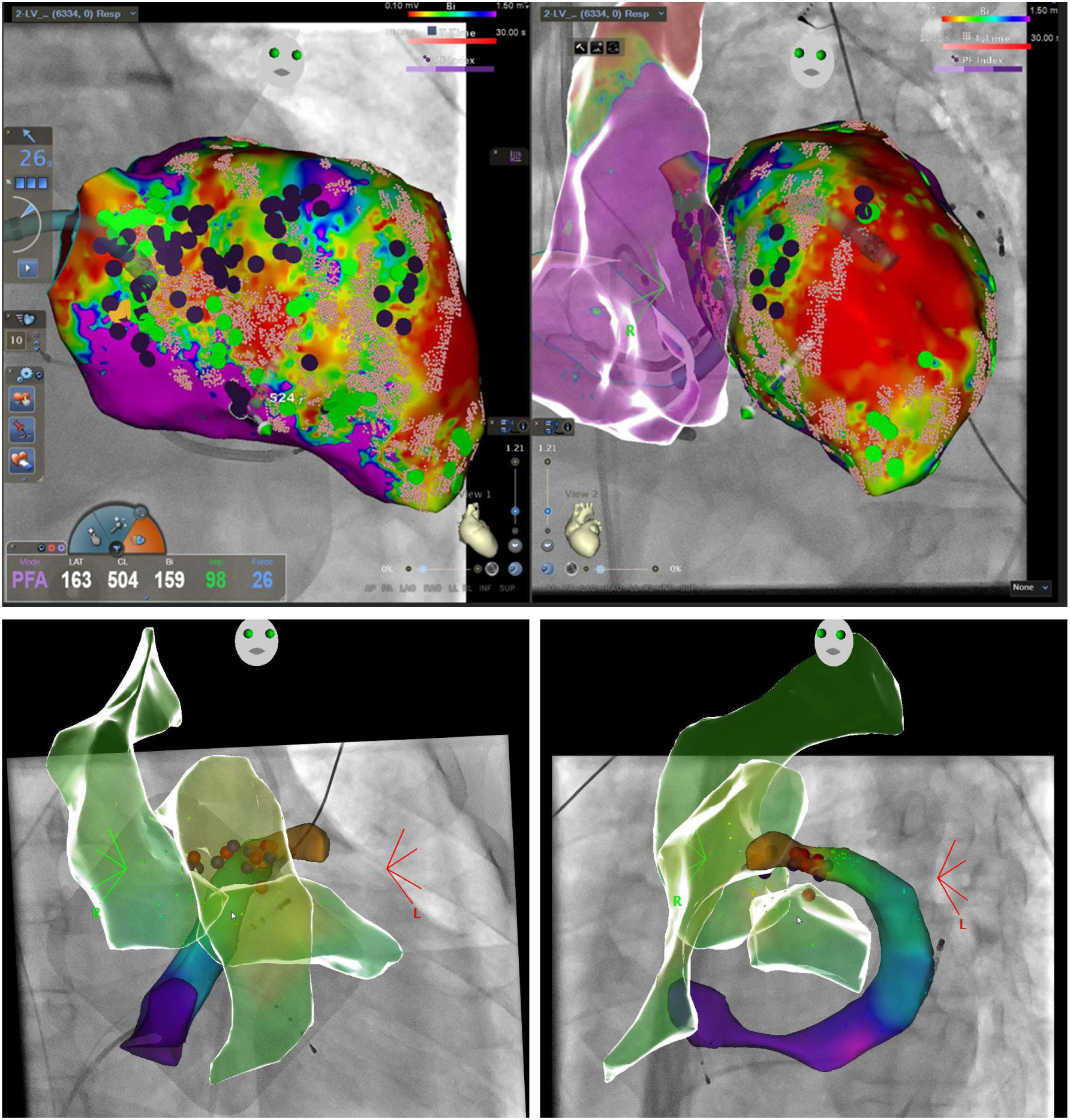
Electroanatomical maps during dual-energy ventricular ablation with the CARTO V8 system. Upper panels: left ventricular bipolar voltage map (0.1–1.5 mV) of a patient with prior anterior myocardial infarction, showing an extensive anterior scar. Green tags indicate late potentials, the pink grid indicates RF lesions, and black tags indicate PF applications. Lower panels: activation map of an LV summit PVC showing earliest activation in the distal coronary sinus, where RF and PF energy were delivered after nitroglycerin administration without coronary spasm. Each map is shown in RAO-LAO projections.

Median procedure duration was 149.5 [120.0–179.8] minutes for VT and 74.5 [59.2–92.0] minutes for PVC ablation, with mapping times of 22.0 [12.0–31.8] and 10.0 [6.8–12.2] minutes and ablation phase durations of 70.5 [52.5–101.5] and 27.5 [20.8–49.2] minutes, respectively. Median fluoroscopy time was 5.9 [4.2–7.0] minutes for VT and 3.0 [1.1–3.9] minutes for PVC procedures, with dose area products of 243.9 [150.2–392.7] and 27.5 [24.3–128.7] cGy·cm² (Table 2). A median of 28.5 [15.0–37.5] PF applications per patient were delivered in VT procedures and 8.5 [7.0–14.8] in PVC procedures. The total energy delivery time, including both RF and PF, was 36.5 [25.2–55.3] minutes in VT and 14.6 [7.6–16.8] minutes in PVC procedures (Tables 2, S1, and S2).

**Table 2.** Procedural characteristics and acute outcomes.

|  | All (n = 26) | VT (n = 18) | PVC (n = 8) |
| --- | --- | --- | --- |
| **Energy delivered** |  |  |  |
| PF applications, n | 18.5 [8.2–35.8] | 28.5 [15.0–37.5] | 8.5 [7.0–14.8] |
| Total energy delivery time (RF and PF), min | 29.6 [14.9–47.8] | 36.5 [25.2–55.3] | 14.6 [7.6–16.8] |
| Successful epicardial access | 3 (11.5) | 3 (16.7) | 0 |
| Procedure duration, min | 120.0 [85.0–169.5] | 149.5 [120.0–179.8] | 74.5 [59.2–92.0] |
| Mapping time, min | 15.0 [10.0–26.2] | 22.0 [12.0–31.8] | 10.0 [6.8–12.2] |
| Ablation phase duration (first to last application), min | 59.0 [30.8–91.0] | 70.5 [52.5–101.5] | 27.5 [20.8–49.2] |
| Fluoroscopy time, min | 4.6 [3.3–6.5] | 5.9 [4.2–7.0] | 3.0 [1.1–3.9] |
| Fluoroscopy dose area product, cGy·cm<sup>2</sup> | 205.5 [81.4–392.7] | 243.9 [150.2–392.7] | 27.5 [24.3–128.7] |
| Acute noninducibility (VT) | — | 16 (88.9) | — |
| Acute PVC elimination | — | — | 8 (100) |
| Major complications | 0 | 0 | 0 |
| Arrhythmia recurrence | 2 (7.7) | 2 (11.1) | 0 (0) |
| Free from recurrence at last follow-up | 24 (92.3) | 16 (88.9) | 8 (100) |

Values are median [interquartile range] or n (%). One PF application denotes one complete delivery cycle represented by one lesion tag. Dose area product was unavailable in 1 VT patient. PF = pulsed field; RF = radiofrequency; other abbreviations as in Table 1.

### Acute efficacy

Programmed ventricular stimulation was performed at the end of all 18 VT procedures. Noninducibility of any sustained VT or VF was achieved in 16 patients (89%). Among the 10 patients presenting with electrical storm or incessant VT, 8 (80%) were noninducible at the end of the procedure. Two patients remained inducible: one with NICM had a fast VT of epicardial origin at the end of the ablation but remained free from recurrence during follow-up, and one with ICM had a VT with inferoapical exit and experienced VT recurrence on the day after ablation. In PVC procedures, acute success was achieved in all 8 patients (100%). The study population, workflow, and principal outcomes are summarized in Figure 2.

**Figure 2.**
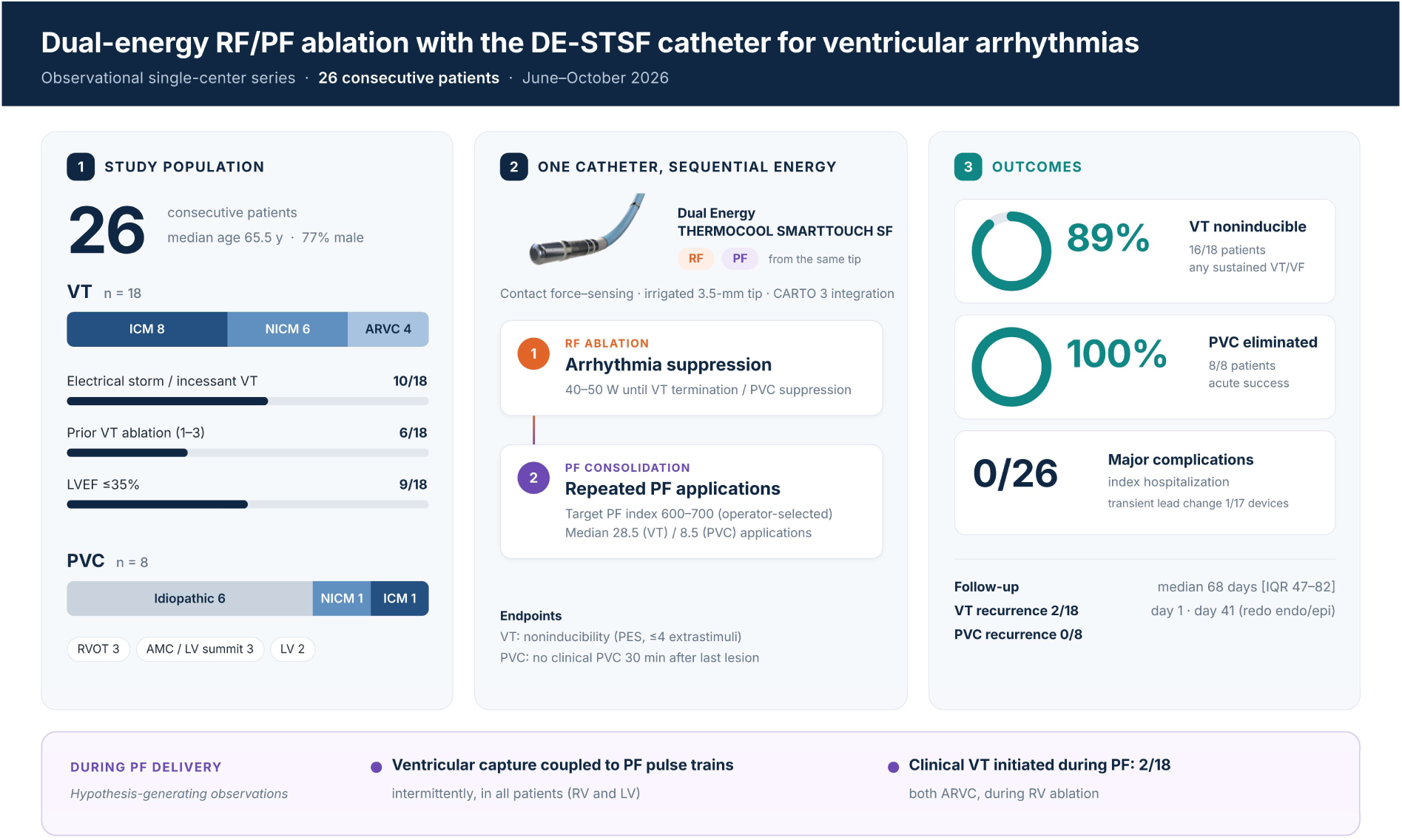
Central illustration. Study population, sequential RF ablation and PF consolidation with the DE-STSF catheter, acute outcomes, early follow-up, and ventricular responses during PF delivery. AMC = aortomitral continuity; ARVC = arrhythmogenic right ventricular cardiomyopathy; DE-STSF = Dual Energy THERMOCOOL SMARTTOUCH SF; ICM/NICM = ischemic/nonischemic cardiomyopathy; LVEF = left ventricular ejection fraction; LVOT/RVOT = left/right ventricular outflow tract; PES = programmed electrical stimulation; PF = pulsed field; PVC = premature ventricular contraction; RF = radiofrequency; VF = ventricular fibrillation; VT = ventricular tachycardia.

### Ventricular responses during PF delivery

Two distinct electrophysiological responses to PF delivery were observed (Figure 3). First, the clinical VT was initiated during PF delivery in 2 of 18 VT patients, both with ARVC, during ablation in the right ventricle. Second, wide ventricular capture during PF applications, with ventricular complexes temporally coupled to PF pulse trains, was observed intermittently in all patients, irrespective of whether energy was delivered in the right or the left ventricle.

**Figure 3.**
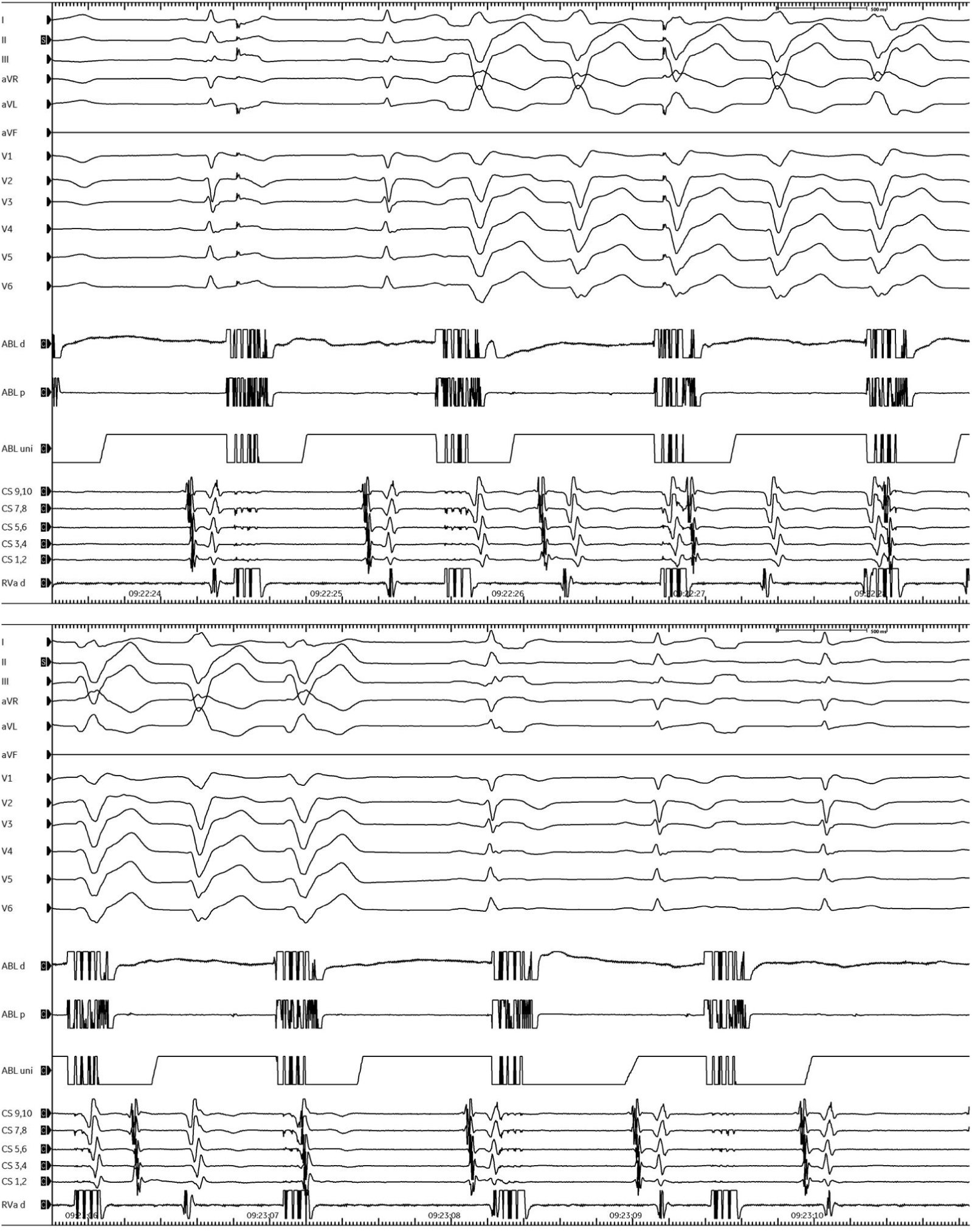
Ventricular responses during PF delivery. Upper tracing: initiation of clinical VT during PF delivery. Lower tracing: ventricular capture associated with PF pulse trains. ABL d/p/uni = distal bipolar/proximal bipolar/unipolar ablation recordings; CS = coronary sinus; PF = pulsed field; RVa d = distal right ventricular apical recording; VT = ventricular tachycardia.

### Safety and early outcomes

No major procedure-related complications were documented in any of the 26 patients during the index hospitalization. There was no clinically recognized coronary spasm, pericardial effusion requiring intervention, stroke, new atrioventricular block, or vascular complication. Seventeen of 18 VT patients (94%) had a CIED (single-chamber ICD in 7, dual-chamber ICD in 5, cardiac resynchronization therapy defibrillator [CRT-D] in 4, and subcutaneous ICD in 1); none of the PVC patients had a device. Devices were manufactured by Medtronic (n = 6), Biotronik (n = 5), Boston Scientific (n = 4), Abbott (n = 1), and MicroPort (n = 1). Post-procedural interrogation showed stable sensing, pacing threshold, and impedance values in 16 of 17 device patients, including those with the highest numbers of PF applications; no generator malfunction or device reset occurred. In one patient with a Biotronik CRT-D, in whom ablation included the basal lateral LV adjacent to the LV lead, post-procedural interrogation showed a decrease in LV lead impedance from 1000 to 400 Ω together with an increase in the pacing threshold of the coronary sinus lead; both parameters had normalized at interrogation on the following day, and no lead revision was required. This was recorded as a device-related observation and did not meet the major complication endpoint. Hemolysis parameters were not systematically assessed (only in 4 of 26 patients and were unremarkable in those tested). As coronary angiography and hemolysis testing were not performed systematically, subclinical coronary spasm and laboratory evidence of hemolysis cannot be excluded.

VT recurrence occurred in 2 of 18 VT patients (11%). The first recurrence occurred on the day after the procedure in an ICM patient who presented with electrical storm, and had a VT with inferoapical exit induced at the end of the procedure. Epicardial access was attempted during the index ablation, but was not successful due to adhesions after bypass surgery. The patient was stabilized after escalation of antiarrhythmic therapy. The second recurrence occurred on day 41 in a patient with ARVC who had presented with electrical storm and had been noninducible at the end of the procedure; the recurrent VT (cycle length 545 ms) was incessant and was treated with antitachycardia pacing and an ICD shock, and the patient subsequently underwent repeat ablation with the DE-STSF catheter using a combined endocardial and epicardial approach. During a median follow-up of 68 [47–82] days, 16 of 18 VT patients (89%) and all 8 PVC patients remained free from recurrence to date.

## Discussion

### Main findings

This study describes the initial clinical experience with a contact force–sensing, irrigated focal catheter capable of delivering both RF and PF energy for the treatment of ventricular arrhythmias. The main findings are as follows: (1) dual-energy ablation was feasible across a heterogeneous spectrum of substrates, including ICM, NICM, ARVC, and idiopathic PVCs from anatomically challenging sites; (2) in a VT population with a high prevalence of electrical storm and prior VT ablation, acute noninducibility of any VT was achieved in 89% of patients; (3) initiation of the clinical VT and ventricular capture were observed during PF delivery; and (4) no major procedure-related complications were documented during hospitalization.

### Rationale for dual-energy ablation in the ventricle

The rationale for combining both energy forms in ventricular ablation rests on their complementary lesion characteristics. RF energy produces well-characterized, resistive-heating lesions whose depth is limited by tissue heating, irrigation, and impedance, and which may be insufficient in thick, septal or scarred myocardium. PF energy, in contrast, is less dependent on thermal conduction and may create homogeneous lesions within fibrotic tissue, with lesion depth increasing with repeated applications.(7) The potential for sequential RF and PF delivery to modify lesion dimensions provides a rationale for further investigation. The principal procedural feature of the platform in the ventricle, however, may not be PF energy itself, but the ability to adapt the energy modality to the procedural step and target within a single catheter and mapping workflow, while retaining contact force information and lesion tagging. In the present series, RF was used for VT termination or PVC suppression, PF for routine consolidation, and RF alone where proximity to a coronary artery was a concern. The RF-first sequence avoided relying on a potentially transient response to PF as the sole acute endpoint. Unlike atrial ablation, in which energy selection is driven mainly by the vulnerability of adjacent extracardiac structures, ventricular energy selection must additionally account for wall thickness, scar heterogeneity, intramural substrate, and the conduction system.

Our cohort included 11 VT patients with septal substrate, 4 patients with ARVC and 3 ablations in the epicardial space, in whom the thin RV free wall, proximity to conduction system, the phrenic nerve and coronary arteries pose specific challenges. These substrates illustrate the range of settings in which this workflow was applied. However, the present data do not establish lesion transmurality, protection of adjacent structures, or the relative contribution of each energy modality.

### Comparison with previous studies

Published experience with ventricular PFA spans early clinical series, feasibility trials, and registry data, with important differences between catheter platforms. Using a high-voltage, force-sensing focal PF catheter, Chiarazzo et al. reported the feasibility of redo VT ablation in patients with NICM.(10) A single-center series of 16 patients with structural heart disease treated with focal PFA reported acute noninducibility as the primary efficacy measure, with septal substrate being a frequent target.(11) First clinical experience with a large-footprint focal catheter toggling between PF and RF energy for VT ablation has also been reported.(12) In the first-in-human VCAS trial, a high-voltage focal PF catheter achieved acute procedural success in 92% of 26 patients with scar-related VT.(23) The multicenter AVAAR registry reported ventricular arrhythmia ablation with a lattice-tip catheter in 126 patients, providing broader clinical experience with another dual-energy platform (13) and CLEAR-VT reported 59 patients, including 50 with VT and 9 with PVCs, treated with Sphere-9. (24) These findings establish a broader clinical context but cannot be transferred directly to the small-tip DE-STSF platform. For DE-STSF specifically, prior reports include an intramural outflow-tract PVC case and sequential RF/PF treatment of incessant post-infarction VT. (16,17) Our series adds catheter-specific procedural data on DE-STSF rather than establishing the feasibility of ventricular dual-energy ablation for the first time. Our acute noninducibility rate of 89% was numerically similar to those reported in these series, although differences in stimulation protocols and patient selection preclude direct comparison. Prospective studies include the randomized ASCEND-VT pilot study and the Sphere-9 VT early feasibility study.(19,20) In atrial fibrillation ablation with a focal dual-energy catheter, the energy modality was selected by target, with PF for the posterior left atrium and RF for the cavotricuspid isthmus or sites near the coronary arteries.(25) By contrast, both energy forms were used in every patient of the present ventricular series, in a fixed sequence, reflecting the different lesion requirements of ventricular substrate.

### VT initiation and ventricular capture during PF delivery

The clinical VT was initiated during PF delivery in two patients with ARVC during right ventricular ablation, while ventricular capture during PF applications occurred intermittently in all patients. Electrical stimulation of excitable myocardium within or surrounding the applied field is a plausible explanation. Experimental and early clinical studies of reversible electroporation demonstrate that subablative pulsed fields can modulate myocardial excitability.(26,27) However, induction of the clinical VT does not establish that the application site is part of the critical reentrant isthmus and should not be considered equivalent to entrainment or confirmation of an appropriate ablation target. Similarly, capture during PF delivery may originate from excitable tissue beyond the irreversible lesion and does not necessarily indicate viable myocardium immediately beneath the catheter tip. Although pacing-based assessment of electrically unexcitable scar and loss of pace capture has been investigated in other ablation settings,(28,29) its relevance to capture during ventricular PF delivery remains unproven. These observations are hypothesis-generating and do not validate a mapping or lesion-completeness endpoint. They also highlight the potential for arrhythmia induction during ventricular PF delivery and the importance of immediate access to pacing and defibrillation.

### Arrhythmia recurrences

One of the two patients who remained inducible at the end of the procedure experienced VT recurrence on the following day and had no further documented VT after escalation of antiarrhythmic therapy. Early recurrences in patients with electrical storm and advanced cardiomyopathy are well recognized after RF ablation and may reflect residual arrhythmogenic substrate and electrical instability. Whether PF lesions differ from RF lesions with respect to early lesion maturation and electrical reconnection in ventricular scar is unknown and warrants dedicated investigation.

A second, later recurrence occurred in a patient with ARVC who had been noninducible at the end of the index procedure. Despite combined RF and PF delivery from the endocardium of the comparatively thin RV wall, residual epicardial substrate sustained a slow reentrant VT (cycle length 545 ms), and stabilization required repeat RF and PF ablation from the epicardium. This case illustrates that endocardial dual-energy delivery, even in thin-walled myocardium, may not achieve transmural lesions across the full thickness of the arrhythmogenic substrate, and that acute noninducibility is an imperfect surrogate of durable substrate modification in ARVC, in which epicardial substrate predominates.

### Safety considerations

No clinically recognized coronary spasm occurred in this series. Coronary spasm is a recognized effect of PF delivery during atrial ablation,(30) and has recently been documented angiographically in 4 of 13 patients undergoing focal ventricular PF ablation near the coronary arteries, frequently without ST-segment changes and associated with a short distance between catheter tip and coronary artery.(31) In the present series, neither coronary angiography nor prophylactic nitroglycerin was used routinely. RF alone was generally used at sites with anticipated proximity to a coronary artery; in two patients with LV summit PVCs, ablation included the distal coronary sinus, and in one of them, combined RF and PF delivery within the distal coronary sinus after administration of nitroglycerin was not associated with coronary spasm (Figure 1). The absence of clinically recognized spasm therefore does not exclude subclinical events, and systematic angiographic assessment should be considered when PF energy is delivered in the outflow tracts, the LV summit, or the coronary venous system.

Patients undergoing VT ablation almost universally carry a CIED, and extensive substrate modification exposes RV and coronary sinus leads to repeated high-voltage PF delivery. Whereas no relevant changes in device function were observed in a small series of patients undergoing PF-based pulmonary vein isolation,(32) MAUDE reports have described generator failure, lead threshold and sensing changes, and program resets after PFA. These malfunctions were reported predominantly in ICD and CRT-D systems and most frequently with the monopolar Affera system and Biotronik devices. (33) In addition, induction of ventricular arrhythmias, as well as transient changes in R-wave amplitude, lead impedance, and pacing threshold with the Sphere-9 catheter has been reported in patients with Biotronik ICD or CRT-D systems. (34,35) In the present series, 17 of 18 VT patients had a CIED, including five Biotronik systems, four CRT-D systems, and one subcutaneous ICD. No generator malfunction or reset occurred, and device parameters remained stable in all but one CRT-D patient. In that patient, decreased LV lead impedance and increased pacing threshold were reported after extensive ablation adjacent to the lead. These normalized within 24 hours without revision, suggesting a transient local effect at the electrode–tissue interface, such as reversible electroporation or edema, rather than structural damage, although a contribution of RF delivery cannot be excluded. These observations support complete device interrogation before and after ventricular dual-energy ablation, with repeat interrogation when abnormalities are detected, and caution when PF energy is delivered close to lead electrodes.

### Limitations

This study has several limitations. First, it is an observational single-center analysis of a small number of patients without a control group, and it reflects the early learning curve with a new system. Second, the cohort is heterogeneous, combining VT and PVC procedures and different substrates, which limits subgroup conclusions. Third, all patients received RF followed by routine PF consolidation, precluding assessment of the incremental contribution of PF. The PF index target was not clinically validated for ventricular DE-STSF ablation. Fourth, follow-up was short, particularly in the most recently treated patients, so that the low number of recurrences during the brief observation period cannot be interpreted as durable efficacy; longer-term data on VT recurrence, ICD therapies, and mortality are required. Finally, safety assessment was limited to clinically documented events during hospitalization; coronary angiography was not performed routinely and hemolysis parameters were available in only 4 patients.

## Conclusions

In this initial clinical experience, sequential RF ablation and PF consolidation using a single contact force–sensing catheter proved feasible for the treatment of ventricular arrhythmias, achieving high acute success with a favorable early safety profile. Whether PF consolidation confers incremental benefit beyond RF ablation alone warrants prospective comparative evaluation.

## Declarations

## Sources of funding

None.

## Disclosures

CS received research support and lecture fees from Medtronic, Abbott, Boston Scientific, and J&J MedTech and is a consultant for Medtronic, Boston Scientific, and J&J MedTech. PS is a member of the advisory boards of Abbott, Boston Scientific, J&J MedTech, and Medtronic. All other authors have nothing to disclose.

## Data availability

The data underlying this article will be shared upon reasonable request to the corresponding author.

## Declaration of generative AI and AI-assisted technologies in the writing process

During the preparation of this work, the authors used Claude (Anthropic) for language editing and manuscript corrections. The authors reviewed and verified all content and take full responsibility for the published article.

## Supplementary material

**Table 1.** Individual characteristics, ablation targets, and outcomes of patients undergoing VT ablation (n = 18)

| # | Substrate | LVEF, % | Storm | Prior abl. | Ablation region | Energy | Non-induc. | VT recurrence | PF applications, n | Energy time, min | Device | Device check |
| --- | --- | --- | --- | --- | --- | --- | --- | --- | --- | --- | --- | --- |
| **Ischemic cardiomyopathy** |  |  |  |  |  |  |  |  |  |  |  |  |
| VT-1 | ICM | 15 | + | 0 | Inferoseptal, inferior, lateral LV (basal to apical) | PF+RF | Yes | – | 23 | 86.1 | VR-ICD | Normal |
| VT-2 | ICM | 20 | – | 0 | LVOT; basal anterolateral, lateral, inferolateral LV; septal RVOT | PF+RF | Yes | – | 36 | 32.5 | CRT-D | Abnormal* |
| VT-3 | ICM | 26 | + | 0 | Apical aneurysm; mid-ventricular anterior to inferior, septal to lateral LV | PF+RF | Yes | – | 38 | 31.9 | VR-ICD | Normal |
| VT-4 | ICM | 30 | – | 0 | Inferior LV (basal to apical, septal to lateral); septal and inferior RV | PF+RF | Yes | – | 18 | 50.3 | VR-ICD | Normal |
| VT-5 | ICM | 45 | + | 0 | Inferoseptal, inferior, inferolateral LV (basal to apical) | PF+RF | No | Day 1 | 39 | 81.4 | DR-ICD | Normal |
| VT-6 | ICM | 50 | – | 3 | Basal inferoseptal, inferior, inferolateral LV | PF+RF | Yes | – | 19 | 33.5 | DR-ICD | Normal |
| VT-7 | ICM | 53 | + | 2 | Basal inferoseptal, inferior, inferolateral LV | PF+RF | Yes | – | 14 | 24.5 | VR-ICD | Normal |
| VT-8 | ICM | 63 | + | 0 | LV apex | PF+RF | Yes | – | 34 | 15.6 | VR-ICD | Normal |
| **Nonischemic cardiomyopathy** |  |  |  |  |  |  |  |  |  |  |  |  |
| VT-9 | NICM | 25 | – | 0 | Anterior, lateral, inferior LV, basal to apical (epi only) | PF+RF | Yes | – | 71 | 11.7 | VR-ICD | Normal |
| VT-10 | NICM | 30 | + | 0 | Diffuse LV, septum, CS, RV (epicardial LV origin) | PF+RF | No | – | 7 | 59.0 | CRT-D | Normal |
| VT-11 | NICM | 30 | – | 0 | AMC; basal anterolateral, anterior, septal, inferoseptal LV | PF+RF | Yes | – | 19 | 27.4 | CRT-D | Normal |
| VT-12 | NICM | 30 | + | 0 | Basal to mid anterolateral to inferolateral LV (endo + epi) | PF+RF | Yes | – | 59 | 45.0 | CRT-D | Normal |
| VT-13 | NICM | 41 | + | 1 | Septal, inferior, lateral basal LV; septal RV | PF+RF | Yes | – | 7 | 48.0 | DR-ICD | Normal |
| VT-14 | NICM | 54 | – | 0 | Basal anteroseptal to anterior LV | PF+RF | Yes | – | 6 | 6.4 | None | n.a. |
| **Arrhythmogenic right ventricular cardiomyopathy** |  |  |  |  |  |  |  |  |  |  |  |  |
| VT-15 | ARVC | 35 | + | 3 | RV free wall, inferior RV, RVOT | PF+RF | Yes | Day 41 | 51 | 96.8 | DR-ICD | Normal |
| VT-16 | ARVC | 43 | – | 2 | RVOT, basal to mid anterolateral RV free wall | PF+RF | Yes | – | 35 | 57.0 | S-ICD | Normal |
| VT-17 | ARVC | 50 | – | 0 | RVOT | PF+RF | Yes | – | 12 | 16.8 | VR-ICD | Normal |
| VT-18 | ARVC | 60 | + | 3 | Basal to mid lateral/inferior RV, RVOT (endo + epi) | PF+RF | Yes | – | 36 | 39.4 | DR-ICD | Normal |

Storm = electrical storm or incessant VT at presentation; VT recurrence = day of first documented recurrence after the index procedure (day 0 = day of the procedure). Device check = complete device interrogation after ablation; \*decrease in LV lead impedance (1000 to 400 $\Omega$ ) and increase in pacing threshold of the coronary sinus lead, normalized on the following day without lead revision. CRT-D = cardiac resynchronization therapy defibrillator; DR-ICD/VR-ICD = dual-/single-chamber ICD; S-ICD = subcutaneous ICD. Energy time = total RF plus PF delivery time. AMC = aortomitral continuity; CS = coronary sinus; LVOT/RVOT = left/right ventricular outflow tract; other abbreviations as in Tables 1 and 2. Non-induc. = noninducibility of any sustained VT or ventricular fibrillation; prior abl. = number of previous ablation procedures; ICM = ischemic cardiomyopathy; NICM = nonischemic cardiomyopathy; LV/RV = left/right ventricle; endo/epi = endocardial/epicardial.

**Table 2.** Individual characteristics, ablation sites, and outcomes of patients undergoing PVC ablation (n = 8)

| # | Substrate | LVEF, % | Prior abl. | PVC origin / ablation site | Energy | Acute success | Recurrence | PF applications, n | Energy time, min |
| --- | --- | --- | --- | --- | --- | --- | --- | --- | --- |
| **Right ventricular outflow tract** |  |  |  |  |  |  |  |  |  |
| PVC-1 | Idiopathic | 60 | 0 | Septal RVOT | PF+RF | Yes | – | 8 | 20.5 |
| PVC-2 | Idiopathic | 55 | 0 | Posterior RVOT | PF+RF | Yes | – | 17 | 4.4 |
| PVC-3 | Idiopathic | 60 | 0 | Anterior RVOT | PF+RF | Yes | – | 14 | 8.3 |
| **Left ventricular outflow tract and LV summit** |  |  |  |  |  |  |  |  |  |
| PVC-4 | Idiopathic | 45 | 0 | AMC, RCC/LCC commissure | PF+RF | Yes | – | 4 | 15.5 |
| PVC-5 | Idiopathic | 40 | 0 | LV summit (LV and distal CS) | PF+RF | Yes | – | 4 | 14.5 |
| PVC-6 | ICM | 38 | 0 | LV summit (distal CS, RVOT, and LVOT) | PF+RF | Yes | – | 9 | 14.6 |
| **Left ventricle** |  |  |  |  |  |  |  |  |  |
| PVC-7 | Idiopathic | 60 | 0 | Anterolateral papillary muscle | PF+RF | Yes | – | 19 | 47.0 |
| PVC-8 | NICM | 53 | 1 | Basal anterolateral LV | PF+RF | Yes | – | 8 | 5.3 |

Patients are grouped by anatomical origin. LCC/RCC = left/right coronary cusp; other abbreviations as in Tables 1, 2, and 3A.

## Notes

### Competing Interest Statement

The authors have declared no competing interest.

### Clinical Trial

observational study

### Author Declarations

Approved by the local ethics committee: Ethics Committee of the Medical Faculty of Ruhr University Bochum, East Westphalia Campus (Ethik-Kommission der Medizinischen Fakult’t der Ruhr-Universit’t Bochum, Sitz Ostwestfalen), Georgstraße 11, 32545 Bad Oeynhausen, Germany.

